# Congenital syphilis outcomes by penicillin dosing intervals among women with late latent or latent of unknown duration syphilis during pregnancy in eight U.S. jurisdictions: 2018-2023

**DOI:** 10.64898/2026.09.03.26361028

**Authors:** Ellen Martinson, Elizabeth L. Lewis, Suzy Newton, Jeffrey M. Carlson, Kathryn Miele, Kevin P. O’Callaghan, Breanne Anderson, Tia Falzarano, Toby R. Levin, Mallory Jayroe, Caroline M. Johnson, Nicole D. Longcore, Adam Berryhill, Teri’ Willabus, Kate R. Woodworth, Van T. Tong

## Abstract

Pregnant women with late latent or latent of unknown duration syphilis are recommended to receive three doses of benzathine penicillin G each 9 or fewer days apart; data on interval variations are limited. We found no significant difference in risk of congenital syphilis between 9-day and 6-8 day dosing intervals.

**Summary:** Among pregnant women with late latent or latent of unknown duration syphilis, there was no difference in risk of congenital syphilis between 9-day and 6-8 day maternal treatment dosing intervals.

## Introduction

Syphilis, a sexually transmitted bacterial infection, is of particular concern during pregnancy due to potential transmission to the fetus, leading to congenital syphilis (CS). Between 2015 and 2024, rates of syphilis increased 405% among women aged 15-44 years,^1^ with CS rates mirroring these increases. Adequate treatment of syphilis during pregnancy can prevent CS and CS-related outcomes, including stillbirth, prematurity, low birth weight, and neonatal intensive care unit (NICU) admission.^2-4^ The 2021 Centers for Disease Control and Prevention (CDC) STI Treatment Guidelines^5^ recommend pregnant women with late latent or latent of unknown duration (LLUD) syphilis receive three doses of benzathine penicillin G (BPG) spaced at one-week intervals with no more than nine days between doses. Few population studies to date have assessed the association between adherence to the one-week dosing interval recommendation and the risk of CS. A recent single-site study reported no difference in the risk of CS among women who received treatment doses on a strict seven-day interval versus those who received doses six to eight days apart^6^; though a preliminary analysis from the same group reported an unadjusted increased risk of CS among those with at least one dosing interval of nine days.^7^ This analysis assesses the risk of CS among pregnant women with LLUD syphilis across eight U.S. jurisdictions by dosing intervals, comparing at least one dose spaced nine days apart and all doses spaced six to eight days apart in a three-dose BPG regimen.

## Materials and Methods

Data included are those reported to CDC as of June 12, 2026 from eight U.S. jurisdictions (Arizona, Arkansas, Georgia, Maricopa County, Michigan, New Jersey, New York [excluding New York City], and Washington), as part of the Surveillance for Emerging Threats to Mothers and Babies Network (SET-NET).^8^ Pregnant woman-infant dyads were included in SET-NET if 1) the pregnant woman met the 2018 Council of State and Territorial Epidemiologists (CSTE) case definition for syphilis at any point during pregnancy or 2) the infant met the probable or confirmed CSTE case definition for CS or met the criteria for syphilis-related stillbirth. The analysis was further restricted to dyads with a reported LLUD maternal surveillance syphilis stage, a reported CS infant case classification, and a reported known pregnancy outcome between January 1, 2018 and December 31, 2023. A reported maternal positive treponemal or nontreponemal test was required for inclusion to confirm syphilis infection during pregnancy. Women pregnant with multiples or who were reinfected during pregnancy were excluded from this analysis, along with those with early pregnancy losses (<20 weeks), terminations, and other unspecified non-live births. For women with repeat pregnancies that met inclusion during the surveillance period, one pregnancy was chosen at random. Methods were consistent with prior studies using this SET-NET cohort.^2^

Dyads were categorized into three exposure groups based on dosing interval: the six-eight day adequate treatment group (three doses of BPG, with each dose spaced six to eight days), the nine-day adequate treatment group (three doses of BPG, with at least one dose spaced nine days and the other doses spaced six to nine days), and the no or inadequate treatment group (treatment initiated less than 30 days prior to the pregnancy outcome, non-BPG treatments, spacing for any dose outside of six to nine days, or less than three doses of BPG). Women who received at least one five-day BPG interval were excluded to align with prior studies.^6,7^

CS was defined as any stillbirth (pregnancy loss at a gestational age >20 weeks), a positive infant direct detection laboratory test (e.g., positive DFA), or a reactive infant nontreponemal serologic test with any of the clinical, laboratory or radiographic findings detailed in the 2018 CSTE surveillance case definition infant-based criteria. Poisson regression was used to calculate unadjusted and adjusted (aRR) risk ratios and 95% confidence intervals (CIs) to assess the association between dosing interval group and CS. Adjusted models accounted for reported substance use during pregnancy, insurance status at delivery, and education. Multiple imputation was conducted to account for missing insurance status at delivery and education to reduce exclusion bias in the final model. Models, including a complete case analysis, are presented in Table 1. Analyses were conducted using R statistical software (4.2.2). This activity was reviewed by the CDC and conducted consistent with applicable federal law and CDC policy (*45 C*.*F*.*R. part 46*.*102(l)(2), 42 U*.*S*.*C. Sect. 241(d); 5 U*.*S*.*C. Sect. 552a*).

**Table 1.** Models to assess association between maternal dosing interval treatment group and congenital syphilis.

| **Treatment group** | **Total (n)** | **Congenital syphilis (CS)<sup>a</sup>**<br>**(n/%)** | **Unadjusted RR (95 %**<br>**CI)** | **Imputed adjusted RR**<br>**(95 % CI)<sup>b,c</sup>** | **Complete case analysis**<br>**adjusted RR (95 % CI)<sup>b,d</sup>** |
| --- | --- | --- | --- | --- | --- |
| All dosing intervals 6-8 days | 836 | 55 (7%) | Reference | Reference | Reference |
| One or more 9-day dosing<br>interval<sup>e</sup> | 41 | 1 (2%) | 0.37 (0.05, 2.61) | 0.36 (0.05, 2.51) | 0.48 (0.07, 3.34) |
| Inadequate/no treatment | 703 | 201 (29%) | 4.35 (3.28, 5.76) | 3.67 (2.74, 4.91) | 3.82 (2.75, 5.30) |

RR: relative risk; CI: confidence interval
<sup>a</sup>For the purposes of this analysis, CS was defined as any stillbirth (pregnancy loss at a gestational age >20 weeks), a positive infant direct detection laboratory test (e.g. positive DFA), or a reactive infant nontreponemal test with any of the clinical, laboratory or radiographic findings detailed in the 2018 CSTE case definition infant-based criteria.
<sup>b</sup>Adjusted risk ratio controlling for reported substance use (alcohol, tobacco, cannabis, illicit opioids, methamphetamines, cocaine, or other illicit non-prescription substances), insurance status at delivery, and education status.
<sup>c</sup>Models imputed for education status (14% missing) and insurance status at delivery (14% missing).
<sup>d</sup>Complete case analysis was restricted to dyads with nonmissing data for all variables included in the adjusted model (n=664 for all dosing intervals of 6–8 days; n=31 for one or more 9-day dosing intervals; n=523 for inadequate/no treatment).
<sup>e</sup>Three doses of benzathine penicillin G, with at least one dose spaced nine days and the other doses spaced six to nine days.

## Results

As of June 12, 2026, 3,388 pregnant woman-infant dyads met SET-NET inclusion criteria. Of those, 1,580 dyads met study inclusion criteria (Figure 1). Nearly half of the included pregnant women were reported as receiving no or inadequate treatment (44% [n=703]), and 56% received adequate treatment, with 836 (95%) women in the six-eight day group and 41 (5%) in the nine-day group (Table 1). The median age of women in the study was 27.9 years. The majority received a high school diploma or less (67%) and had public insurance (65%). Among women who received no or inadequate treatment, 63% reported substance use compared to 34% in the six-eight day group and 46% in the nine-day group.

**Figure 1:**
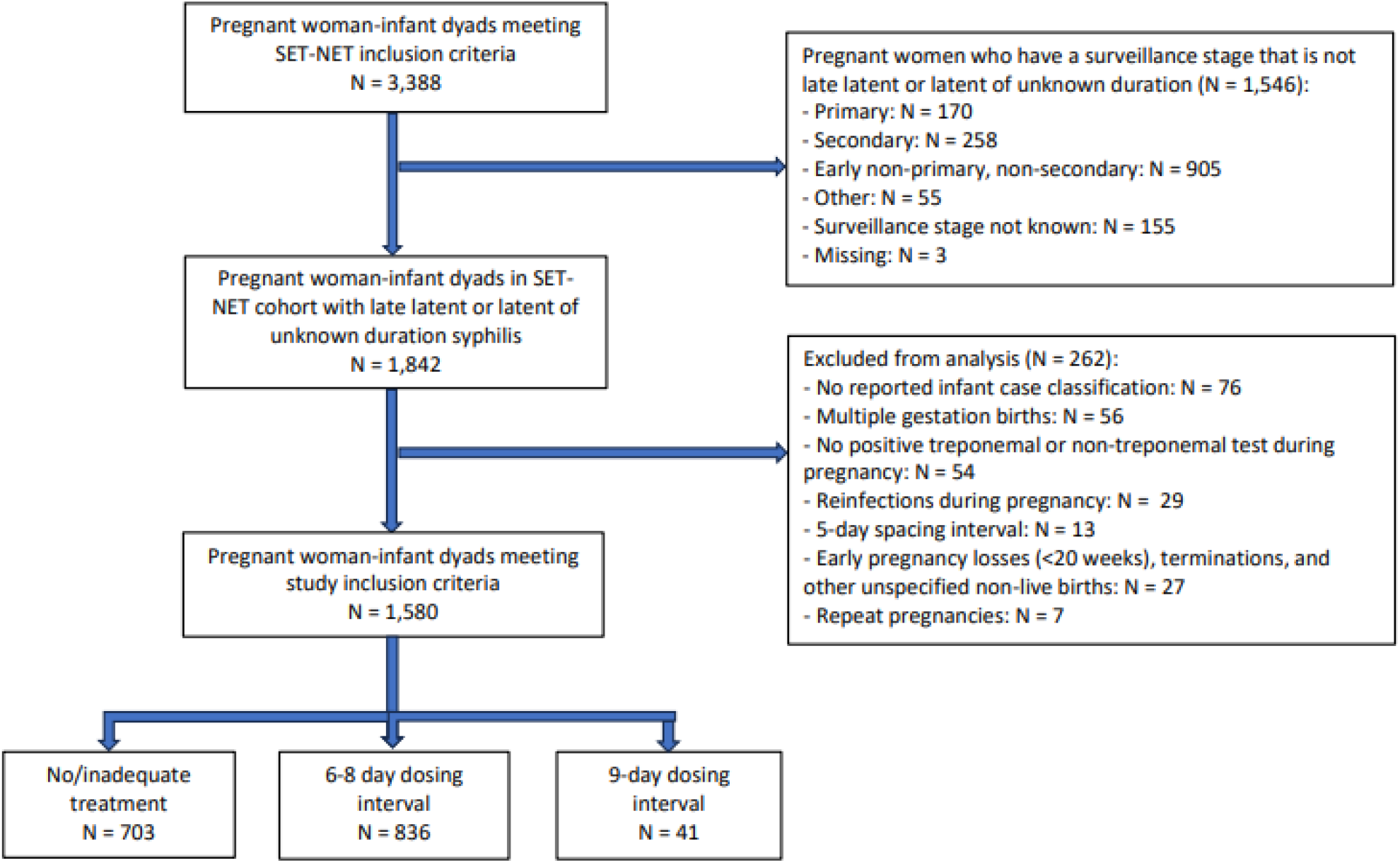
Study inclusion and exclusion flowchart of pregnant woman-infant dyads from SET-NET cohort, reported as of June 2026.

The frequency of CS was 29% among infants born to women in the no or inadequate treatment group, 7% in the six-eight day group, and 2% in the nine-day group (Table 1). The adjusted risk of CS among infants born to women in the nine-day group was 0.36 [95% CI: 0.05, 2.51] compared to the six-eight day group (Table 1). Infants born to women who received no or inadequate treatment had a significantly elevated risk of CS (aRR: 3.67 [95% CI: 2.74, 4.91]) compared to the six-eight day group. All risk estimates were in the same direction and of similar magnitude in the complete case analysis. The proportions of infants who were small-for-gestational-age, admitted to the NICU, or born preterm were not statistically significantly different between the six-eight day and nine-day groups.

## Discussion

We did not detect a statistically significant association between risk of CS among infants born to women with LLUD syphilis and maternal treatment with at least one BPG spacing interval of nine days compared to all spacing intervals between six and eight days, though the small number of women in the nine-day group limits interpretation. No statistically significant differences in small-for-gestational-age, NICU admission, or preterm birth between the six-eight day and nine-day groups were observed.

The strength of this study is its large, multi-state, multi-year cohort that allowed us to classify CS among all infants exposed to syphilis during pregnancy based on reported clinical and laboratory findings rather than solely infants who met the CSTE case definition for confirmed or probable CS. The only study assessing the association between a nine-day interval and risk of CS showed an elevated, unadjusted risk of CS of 3.7 in the nine-day group compared to the six-eight day group.^7^ Our results do not demonstrate the same elevated risk of CS for infants born to women in the nine-day group compared to the six-eight day group. The recommendation in the 2021 STI Treatment Guidelines is based on a single, small (n=25) pharmacokinetic study of pregnant women receiving medication just before delivery which found non-significant decreases in serum penicillin G concentration between days one and seven post administration.^9^ Though interpretation should be limited due to a small number in the nine-day group, our study results support the 2021 Treatment Guidelines that permit a nine-day BPG treatment spacing without restarting the treatment course. Defining acceptable dosing intervals is critical for determining whether a pregnant woman restarts treatment, which is especially important due to ongoing national BPG shortages^10^ and barriers to accessing and accepting treatment.^11^ Flexible dosing intervals may also reduce unnecessary work up of infants.^5^

Our study has the following limitations. CS was defined using reported clinical and laboratory findings. Since SET-NET relies on medical record abstraction, underreporting of these findings due to missing records might underestimate the true prevalence of CS in our sample. Treatment may also be underreported due to surveillance limitations, leading to exposure misclassification towards the no or inadequate treatment group, for example if treatment dates are only reported to SET-NET for two of the three doses. Additionally, maternal neurosyphilis was not systematically collected in SET-NET prior to 2022 births (n=4; 2022-2023) and were therefore not excluded from analysis. Since the treatment regimen differs for neurosyphilis, these dyads might be misclassified in the inadequate treatment group. Finally, these findings may not be generalizable to jurisdictions or years outside the included sample.

No significant difference was observed in risk of CS for infants born to women with LLUD syphilis treated with at least one dose of BPG spaced at nine days compared to all doses within the six-eight day range. These findings support the 2021 STI guidelines that permit more flexible dosing intervals of up to nine days. By allowing a more flexible window for prenatal treatment dosing and studying the effects of variable dosing intervals, we can improve prenatal syphilis treatment and reduce CS in the U.S. while responsibly managing antibiotics and resources.

## Data Availability

These data are collected under relevant provisions of the Public Health Service Act and are protected at CDC by an Assurance of Confidentiality (Section 308(d) of the Public Health Service Act, 42 U.S.C. 242 m(d))(https://www.cdc.gov/od/science/integrity/confidentiality/), which prohibits use or disclosure of any identifiable or potentially identifiable information collected under the Assurance for purposes other than those set out in the Assurance. Requests for access will be considered on a case by case basis, and inquiries should be directed to.

## Acknowledgements

We thank the staff supporting the SET-NET work, including the Michigan Department of Health & Human Services, Bureau of HIV/STI Programs, Cashea Anderson, Arnelle Anekwe, Nicole Coppola, Deborah Gleissner, Dana Higgins, I-Zis Kerr, Valerie Piccarillo, Mark Rogers, Abigael Gardere, Dyeshia Leonard, and Danielle Rolley of the New Jersey Department of Health, Emily M. Bruce, Nadia Thomas, Ciarra McFarland, and Kaylee Mahoney of the New York State AIDS Institute Office of Sexual Health and Epidemiology, and Kimberly Bryant, Makala Davis, Jonathan Bell, Sabrina Sanchez, and Stephanie Devlin of the Maricopa County Department of Public Health for data collection, reporting, and partnership in this important work.

## Appendix 1. Characteristics of women with late latent or latent of unknown duration syphilis and their neonates.

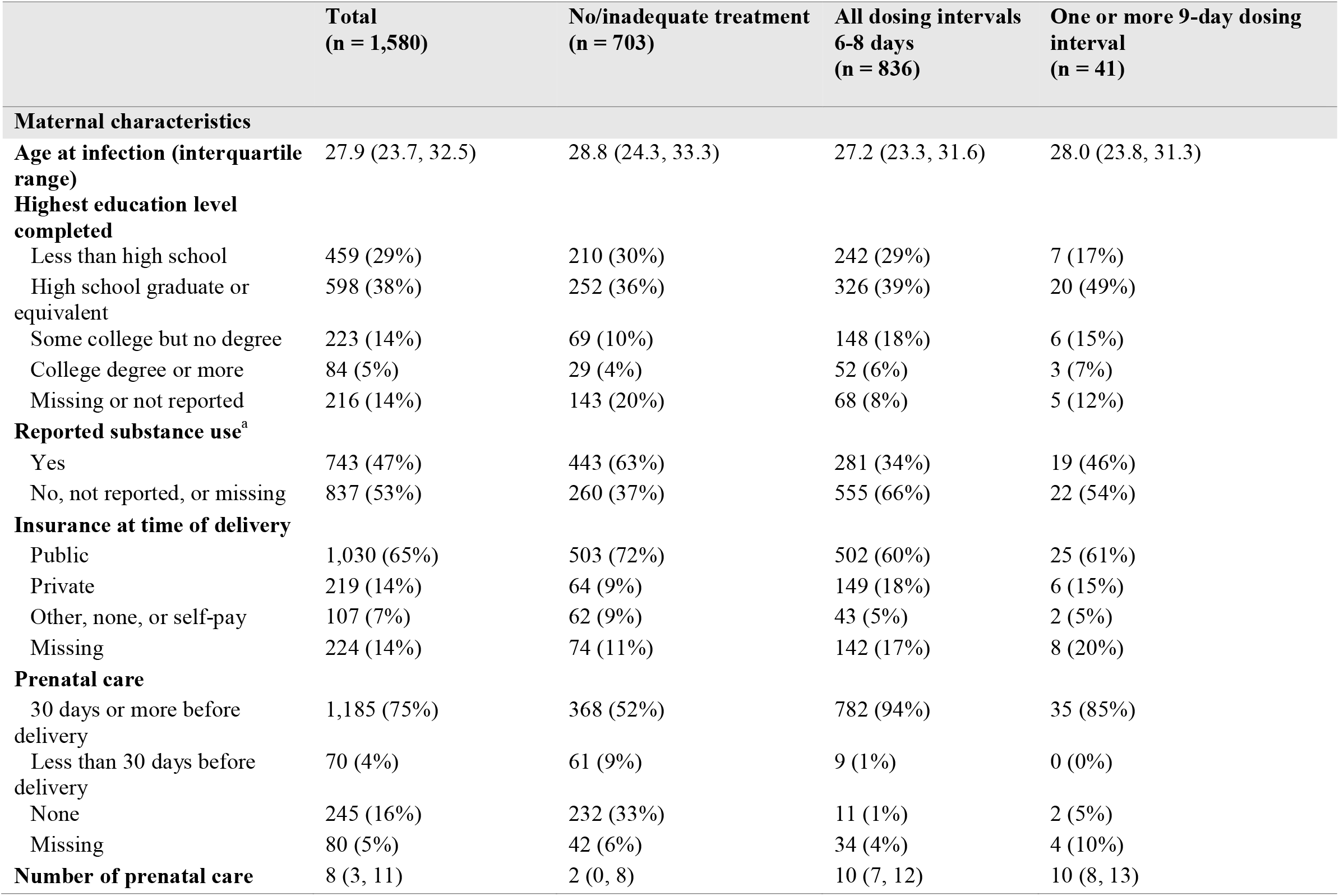
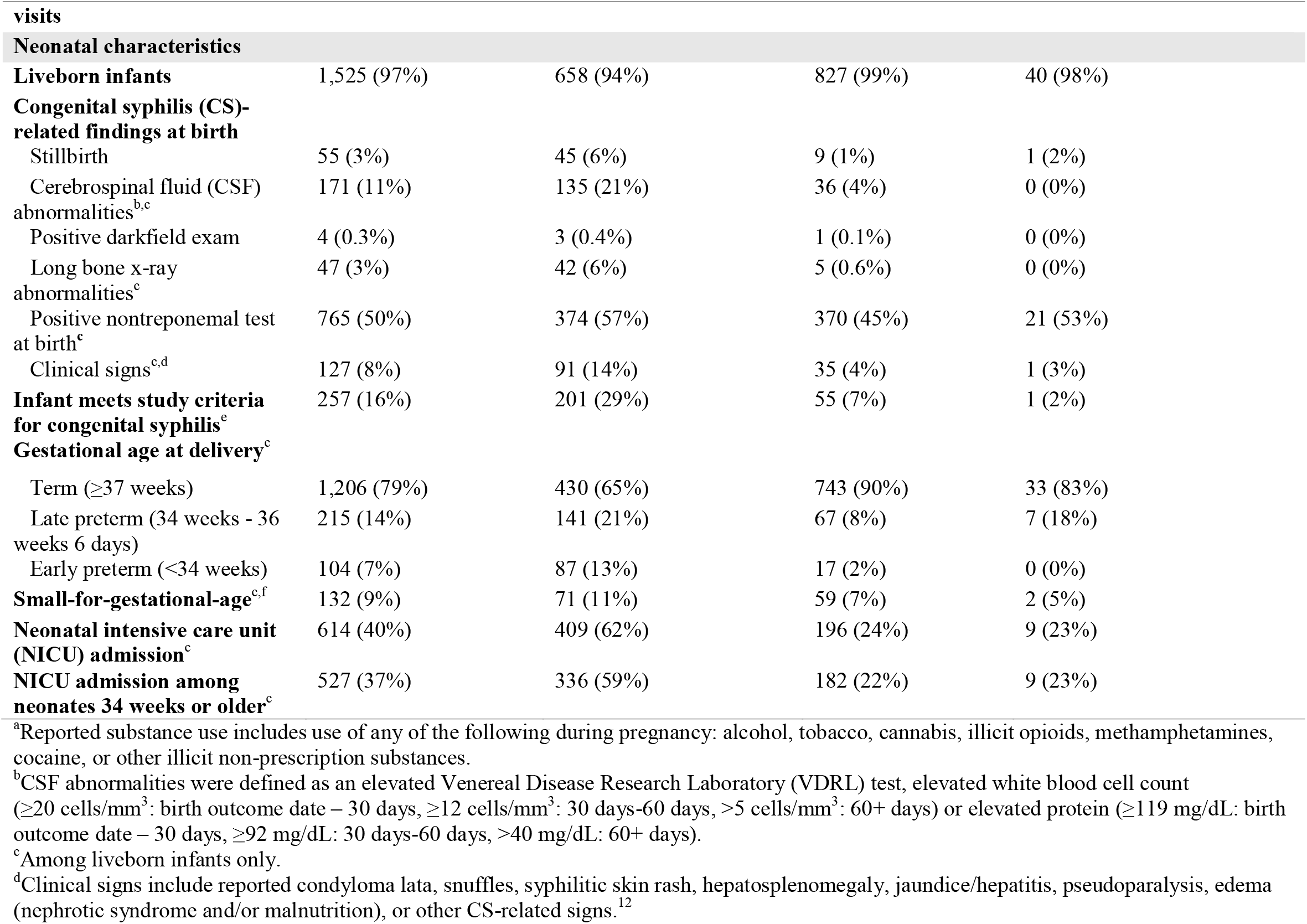
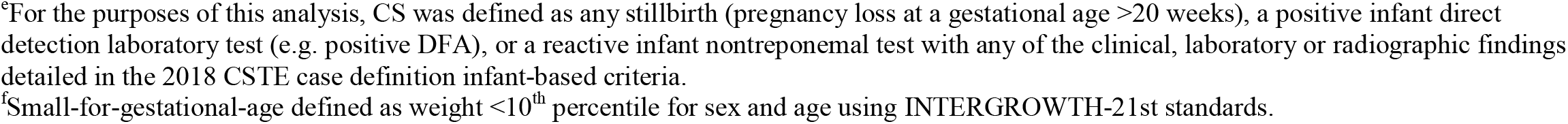

